# Targeting Multiple Immune Checkpoints with a Single Therapy: Implications for Treating Central Nervous System Tumors

**DOI:** 10.64898/2026.02.10.26345679

**Authors:** Meghna Saxena, Elisabeth Ampudia-Mesias, Sumant Dhawan, Stephen C. Frederico, Xiaoqing Cheng, Elisabeth Neil, Ron Bose, Gary Kohanbash, Christopher L Moertel, Michael R Olin

## Abstract

**Background:** Immune checkpoint inhibition has transformed cancer therapy; however, many patients fail to respond to single-agent blockade, and combination strategies are often limited by toxicity. Central nervous system tumors exploit multiple immunosuppressive pathways, including the CD200 and PD-1/PD-L1 axis to evade anti-tumor immunity and support tumor aggressiveness.

**Methods:** We investigated ARL200, a peptide ligand targeting the CD200 activation receptor (CD200AR) using in vitro immune assays, murine syngeneic tumor models, phosphoproteomics, and correlative studies from a first-in-human trial in recurrent glioblastoma.

**Results:** ARL200 exposure activated DAP10/12-dependent signaling and downregulated multiple inhibitory immune checkpoint receptors, including CD200R1, PD-1, and CTLA-4, and checkpoint ligands, CD200 protein and PD-L1, through suppression of the JAK1/3–SHP–STAT–IKKα/β–NFκB pathway. Distinct ARL200 variant peptides elicited unique immune responses. In patients with recurrent glioblastoma, ARL200 treatment was associated with immune activation, reduced inhibitory checkpoint expression, and evidence of antigen-specific memory responses without treatment-related toxicity.

**Conclusions:** Targeting CD200AR enables coordinated modulation of multiple immune checkpoints with a single agent, representing a next-generation immunotherapeutic strategy opening a new pathway for treating aggressive malignancies.

**Key Points:**

- ARL200 elicits an active immune response for the development of a potent and durable anti-tumor response
- ARL200 abolishes the suppressive effects of multiple immune checkpoint blockades
- Different ARL200 sequences drive alternative immune responses.

**Importance of the Study:** Tumors exploit multiple immune checkpoint pathways to suppress antitumor immunity, particularly within the immunosuppressive microenvironment of the central nervous system. Current immune checkpoint inhibitors often require combination therapy to achieve clinical efficacy, frequently at the cost of increased toxicity. In this study, we demonstrate that targeting the CD200 activation receptor (CD200AR) with a peptide ligand provides a novel strategy to simultaneously downregulate multiple inhibitory immune checkpoints, including CD200R1, PD-1, PD-L1, and CTLA-4, through a shared intracellular signaling pathway. ARL200 engagement activates DAP10/12-dependent signaling while suppressing the JAK1/3–SHP–STAT–IKKα/β–NFκB axis, thereby overriding tumor-mediated immunosuppression. Importantly, this multi-checkpoint modulation is achieved with a single therapeutic agent and translates to immune activation and clinical responses in patients with recurrent glioblastoma, with minimal treatment-related toxicity. These findings establish CD200AR targeting as a next-generation immunotherapeutic approach with the potential to improve the safety and efficacy of immune-based therapies for aggressive CNS malignancies.

**Graphical Abstract:** 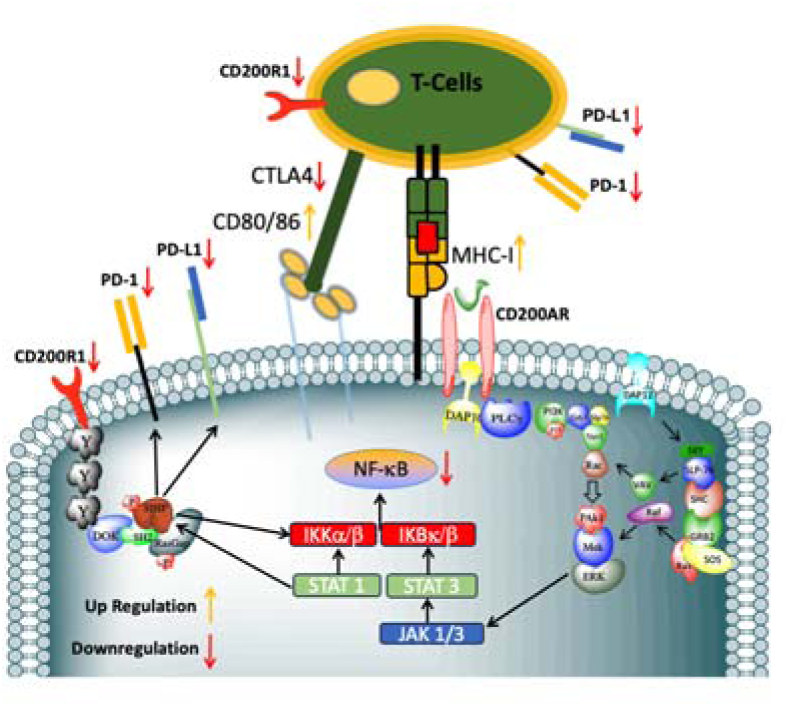

## Introduction

Arming the immune system to mount a robust anti-tumor response is an attractive therapeutic approach for primary and metastatic brain tumors, as the cytotoxic and memory responses of its effector cell populations can be targeted in the tumor microenvironment.^1, 2^ Immune responses are modulated via immunological checkpoints that are often utilized by tumors to evade immune surveillance, and are critical in maintaining homeostasis and protective immunity.^3^

Targeting co-inhibitory or co-stimulatory receptors tightly control T-cell activity preventing unwanted immune responses or enhancing T-cell activation.^3, 4^ Using immunotherapy to block negative checkpoint regulators (NCR) to T-cells leads to an immune activity in the tumor microenvironment.^5, 6^ At the forefront of immunotherapy development are immune-checkpoint inhibitors, which have seen unparalleled success in cancer therapy due to their broad bioactivity across many tumor types. Among the immune checkpoint-blocking strategies, the two most successful to date are those targeting cytotoxic T-lymphocyte-associated protein 4 (**CTLA-4**) and of the interaction between programmed cell death-1 (**PD-1**) and its ligand (**PD-L1**).^4, 6^

The blockade of NCRs with current FDA-approved checkpoint inhibitors is intensely pursued as a therapeutic target for cancer, leading to superior clinical responses in selected solid tumors, however, has little to no response against central nervous system tumors is observed.^7, 8^ Blocking inhibitory checkpoints with agonist monoclonal antibodies against CTLA-4, PD-1 and other inhibitory receptors appears critical for maintaining self-tolerance and modulating immune responses.^9, 10^ This has proved useful in boosting the activity of CD8^+^ cytolytic T-cell activity in tumor immunotherapy. However, only a fraction of cancer patients benefit from the use of a single checkpoint inhibitor.^11^ Although blockade of both the PD-1 and CTLA-4 pathways is important to achieve clinical efficacy, the use of multiple checkpoint inhibitors often intensifies adverse effects, including mortality.^12^ ^13–15^

To address this critical unmet medical need, we developed a unique immunotherapeutic agent designed to enhance anti-tumor immunity by actively modulating the immune system through a series of signaling pathways, opening the door to new methods of treating cancer.^16–20^ One such pathway, the CD200 immune checkpoint, is negatively regulated by the ligation of the inhibitory receptor (CD200R1) and its ligand CD200 protein, which is highly expressed throughout the central nervous system and plays a pro-tumorigenic role.^21, 22^

In our murine syngeneic CD200 knockout glioma model, we demonstrated that CD200 knockout tumors were spontaneously rejected.^23^ This was dependent on a functional immune system, as mice void of natural killer and CD8 T-cells succumb to disease.^23^

Rigorous studies, including our own, suggest that targeting the CD200 pathway is important for the fight against cancer.^24–27, 23^ The CD200 immune checkpoint modulates immune responses through paired receptors.^21, 28^ While others have characterized the CD200 protein and the CD200R1,^21, 28^ we are the first to investigate the mechanism of the CD200 activation receptors (CD200AR), which are predominantly expressed on immune cells, both of myeloid and lymphoid origin.^16, 18–20^ Given the immunostimulatory capacity of CD200AR and the lack of known CD200AR ligands, we developed a novel peptide, ARL200 (formerly known as CD200AR-L).^17, 20^ ARL200 binds to CD200AR, driving two separate mechanisms for a durable anti-tumor response. The first mechanism is that ARL200-CD200AR ligation enhances cytokine/chemokine production, driving myeloid cell differentiation into dendritic cells. This enhances co-stimulation through CD80/86 expression and enhances antigen presentation.^16, 18, 19^ These events are all required for an active T-cell response. A second mechanism through which ARL200/CD200AR ligation activates an immune response is through DNAX-activating 10&12 (DAP10&12).^27^ Following our discovery of DAP10 and DAP12 pathway involvement, we initiated studies to further elucidate downstream signaling and the connections between the CD200 and PD-1, CTLA4, and PD-L1 pathways. Here we report that ARL200-CD200AR ligation signals through a unique pathway driven by DAP10&12-JAK1/3-STAT1/2-SHIP-IKKα/β-NFκB, downregulating the inhibitory receptors CD200R1, PD-1, and CTLA-4 and their ligands CD200 protein and PD-L1. In addition, we showed that targeting the CD200AR with different ARL200 peptides drives unique immune responses, which appear to be driven through alternative signaling pathways.^16, 19, 20^

## Methods

### Cell isolation and stimulation

Wildtype mice were splenectomized, and spleens were used to isolate the cells. CD11b and T cells were isolated using microbeads following the manual instructions. Briefly, cells were magnetically labeled with CD11b or CD3 MicroBeads, (Miltenyi Biotec, Auburn, California United States), and then cell suspension was loaded onto a MACS column (Miltenyi Biotec, Auburn, California United States), which was placed in the magnetic field of a MACS separator (Miltenyi Biotec, Auburn, California United States). Afterward, two million cells were seeded in 200ul RPMI media (Corning, Corning, New York) in a 48-well plate and let them rest for 12 hours. Then cells were washed with 1X PBS and pulsed with 10uM ligand P1 for one hour. Non-Pulsed and LPS-pulsed cells were used as negative and positive controls respectively.

#### qPCR studies

Total RNA was extracted using TRIzol (Thermo Fisher, Waltham, MA) following the manufacturer’s instructions. qPCR was carried out using specific primers for for glyceraldehyde-3-phosphate dehydrogenase (mGAPDH), programmed cell death protein 1 (mPD1), CD200 receptor 1 (mCD200R1). mGAPDH, mP38MAPK, mERK1/2, mSLP76, mPIk3, mcJUN, mJAK1, and mJAK2 as described in supplementary methods.

### CRISPR–Cas9–mediated deletion of CD200R1 in Raw 264.7 macrophages

Raw 264.7 macrophages were maintained in RPMI 1640 supplemented with 10% fetal calf serum (FCS) and 1% penicillin/streptomycin at 37°C and 5% CO_₂_ until ∼80% confluency. Cells were harvested by trypsinization, washed once with RPMI, and resuspended in PBS at a density of 5 × 10 cells/mL. For each electroporation, 5 × 10 cells (10 µL) were pelleted and resuspended in 10 µL Neon Buffer R.

Cells were mixed with 1 µg CleanCap Cas9 mRNA (TriLink Biotechnologies) and 100pmol CRISPRevolution sgRNA targeting CD200R1 (Synthego) and incubated for 2 min at room temperature. Electroporation was performed using the Neon Transfection System (Thermo Fisher Scientific) with the following settings: 1,720 V, 10 ms pulse width, and 2 pulses. Following electroporation, cells were immediately transferred to complete medium and cultured for 48 h.

Genomic deletion of CD200R1 was confirmed by PCR amplification and sequencing of the targeted locus. Loss of CD200R1 surface expression was assessed by flow cytometry using PE-conjugated anti-CD200R1 antibody (OX-110; BioLegend).

#### Cytokine Secretion

A total of 1 × 10^6^ cells was grown in 0. 500 ml in RPMI-1640 in a 48-well plate for 12 h, then pulsed with 10uM ARL200, and incubated for an additional 48 h. Supernatant (50ul) were collected and analyzed for TNFα levels using cytometry bead array (Biosciences). Data was analyzed using Flowjo v10.

#### Western Analyses

Cells were stimulated with mCD200AR-L for (0, 1, 3, 5, 7, 10, 15, 30, 60 min). Following stimulation, cells were lysed ana analyzed by western analysis for CD200R1, CD200R2, CD200R3, CD200AR4, PD-L1, PD-1 and PDL1 as described in supplementary methods.

#### Fe-IMAC Sample preparation and LC-MS/MS analysis

CD11b cells were pulsed with 10 μM CD200AR-L, cellular extracts were sent to Cell Signaling Technology for PTMScan Fe-IMAC analysis. Extracts were sonicated, centrifuged, reduced with DTT, and alkylated with iodoacetamide. 500 μg total protein for each sample was digested with trypsin, purified over C18 columns and used for IMAC enrichment using Fe-NTA magnetic beads (CST #20432) as previously described ^29^. LC-MS/MS analysis was performed using an Orbitrap-Fusion Lumos Tribrid Mass spectrometer as previously described ^29, 30^ with replicate injections of each sample for the IMAC analysis. Briefly, peptides were separated using a 50cm x 100µM PicoFrit capillary column packed with C18 reversed-phase resin and eluted with a 150-minute linear gradient of acetonitrile in 0.125% formic acid delivered at 280 nl/min.

MS/MS spectra were evaluated by Cell Signaling Technology using SEQUEST and the GFY-Core platform (Harvard University, Cambridge, MA). Searches were performed against the most recent update of the Uniprot *Mus musculus* database with a mass accuracy of +/- 50ppm for precursor ions and 0.02Da for-product ions. Results were filtered to a 1% peptide-level FDR with mass accuracy +/- 5ppm on precursor ions and presence of a phosphorylated residue for IMAC enriched samples. Results were further filtered to a 1% protein level false discovery rate. All IMAC quantitative results were generated using Skyline^31^ to extract the integrated peak area of the corresponding peptide assignments. Accuracy of quantitative data was ensured by manual review in Skyline or in the ion chromatogram files.

#### Statistical analysis

Sample sizes were selected based on data from previous experiments done in our laboratories and published results from the literature. Animal experiments were performed after randomization. To estimate statistical significance of differences between 2 treatment groups, a two-tailed *P*-value was calculated using an unpaired *t*. P values less than 0.05 were considered statistically significant. Statistically significant *P*-values were indicated in the figures as follows: **** *P<.001,* \*\*\**P* < .001, \*\**P* < .01, and \**P* < .05. Data were analyzed using Prism software v. 6.01 (GraphPad).

#### Ethics Statement

All animal studies were conducted in accordance with institutional guidelines and were approved by the Institutional Animal Care and Use Committee (IACUC). All human subject data included in this manuscript were obtained in compliance with established Institutional Review Board (IRB) protocols, and all applicable ethical standards and regulations were followed. IRB of University of Minnesota gave ethical approval for this work.

## Results

### ARL200 downregulates CD200R1, PD-1, and PD-L1 expression *in vitro* and *in vivo*

The CD200/CD200R1 axis serves a pivotal role in maintaining immune tolerance and protecting healthy tissues from immune damage. It is implicated in the pathogenesis of solid tumors and hematologic malignancies through the mediation of innate and adaptive immunosuppression. Evidence from prior studies, including our own, suggests that antitumor immunity can be enhanced by targeting the inhibitory CD200–CD200R1 complex.^25–27, 32, 33, 23^

Our focus has been on the CD200AR, predominantly expressed on immune cells, both of myeloid and lymphoid origin.^9^ Unlike CD200R1, the activation receptors have short cytoplasmic regions lacking independent signaling capacity and require adapter proteins (such as DAP12, DAP10, CD3ζ, and FcRγ) to facilitate signaling through Src homology 2 domains,^8^ and are made up of multiple activation receptor complexes.^20^ We developed a set of peptide inhibitors (ARL200) to target these activation complexes. We reported that intradermal injection of ARL200 activates local immune cells *via* DAP10/12 signaling, boosting chemokine and cytokine production, which recruits other immune cells to the injection site.^20^ Interestingly, the time course of the ARL200 effect is biphasic, with an upregulation of DAP10 & DAP12 noted 24 hours post-stimulation. In these studies, the same effect was observed with each different activation receptor (murine CD200AR2, CD200AR3, and CD200AR4) **(Figure 1a)**. However, in contrast to the CD200ARs, CD200R1, PD-1, and PD-L1 all remained downregulated 24 hours post-stimulation **(Figure 1b)**. To follow up, CD11b cells were pulsed in vitro with ARL200 and analyzed for protein levels of CD200R1, PD-1, and PD-L1 24 hours post-stimulation which demonstrated that the ARL200 downregulated the expression of all three inhibitory molecules. (**Figure 1c)**.

**Figure 1.**
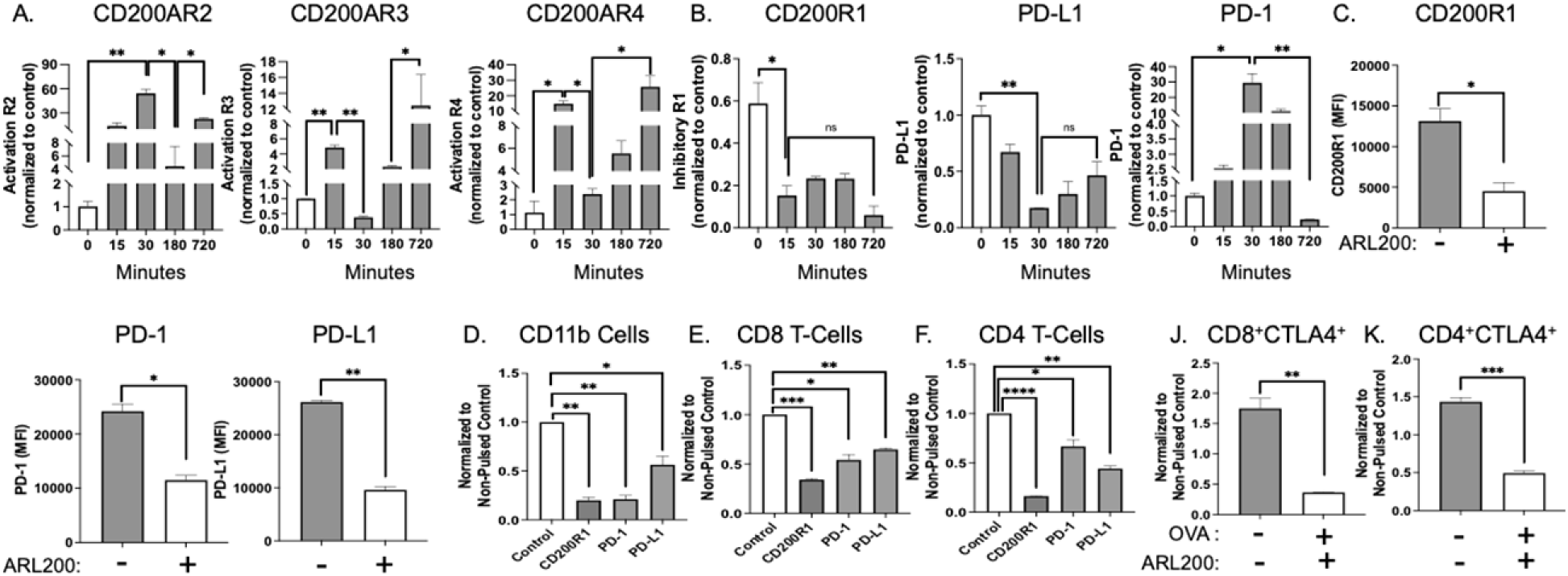
ARL200 inhibits immune checkpoints molecules. CD11b^+^ myeloid cells isolated from WT mice were pulsed with 10 µM (0.2 µg) ARL200. Transcription levels of **(A)** CD200AR2, CD200AR3, **&** CD200AR4, and **(B)** CD200R1, PD-1, and PD-L1 were assessed by RT-PCR between 0- and 720-min. Data were normalized to GAPDH. (**C-E**) CD11b^+^ myeloid cells pulsed with 10 µM ARL200 were analyzed for protein levels of CD200R1, PD-1, and PD-L1. Mice were subcutaneously inoculated with ARL200 above the inguinal lymph node (LN) daily for 5 days. **(D)** CD11b cells **(E)** CD8 T-cells and **(F)** CD4 T-cells were isolated from the inguinal lymph nodes and analyzed for CD200R1, PD-1, and PD-L1 expression by flow cytometry. Mice were subcutaneous inoculated with ARL200 + OVA above the inguinal lymph node daily for 5 days. **(G)** CD8 T cells and **(H)** were assessed for expression of CTLA-4 by flow cytometry. Data are relative to positive stain. Error bars, SD; *p < 0.05; **p < 0.01 by multiple comparison test. Representative data from 3 experiments with n=3 /group.

Next, we sought to determine if ARL200 downregulated the inhibitory molecules in vivo. To accomplish this, 10μM ARL200 was administered intradermally to non-tumor bearing mice for 5 consecutive days. Inguinal lymph nodes were then isolated and CD11b, CD4 T-cells, and CD8 T-cells were analyzed for protein expression of CD200R1, PD-1, and PD-L1 **(Figures 1d-f)**. To investigate the effect of ARL200 on CTLA-4, mice were stimulated with OVA_257-264_ prior to ARL200 as described above. We observed CTLA-4 was downregulated on both CD4 T-cells and CD8 T-cells isolated from the inguinal lymph nodes following administration of ARL200 **(Figures 1g&h)**. These experiments demonstrate that the ARL200 can modulate multiple immune checkpoint molecules in vivo.

### The inhibitory CD200 and PD-1 pathways crosstalk, allowing ARL200 to override the suppressive effects of the CD200 and PD-L1 ligands

Similar to PD-1 and CTLA-4, the CD200 immune checkpoint signals through SHP1/2, connecting them with the PI3K-Akt and Ras-MEK-ERK signaling pathways.^20^ In addition, the inhibitory CD200 protein is upregulated along with PD-L1 and the PD-1 pathway^34^ ^35^ through the use of common signaling molecules, linking them for the development of an immunosuppressive environment.^36^ In a new set of experiments, we sought to determine if an interconnection exists between the inhibitory receptor CD200R1 and the PD-1 pathway. To accomplish this, we knocked out the CD200R1 gene and confirmed the loss of SHIP phosphorylation (**Figure 2a**). We then stimulated wild-type and CD200R1KO macrophages with phorbol 12-myristate 13-acetate (PMA), a mitogen to stimulate cytokine production +/- PD-L1. Following a 48-hour incubation, supernatants were analyzed for TNFα. We observed that the addition of PD-L1 in wild-type cells induced a suppressive response, measured by the decrease of TNFα levels. In contrast, knocking out the inhibitory CD200R1 pathway inhibited the suppressive effects of PD-L1, suggesting that the CD200R1 and PD-1 pathways are connected.

**Figure 2.**
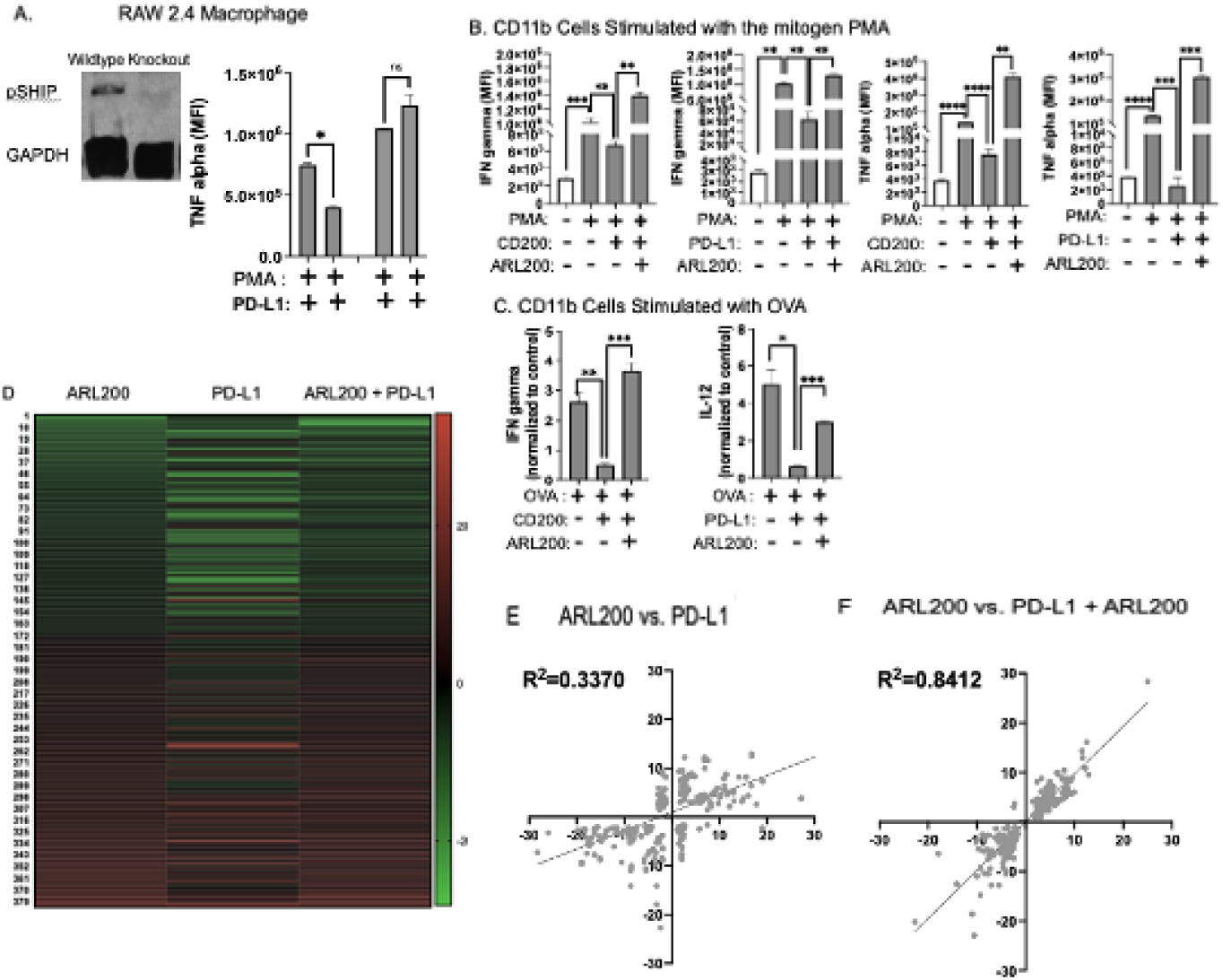
Effect of PD-L1 is inhibited by knocking out CD200R1. **(A)** WT or CD200R1-KO macrophages were pulsed with PD-L1 and analyzed for pSHIP; WT or CD200R1-KO cells were pulsed with PMA ± PD-L1 and analyzed for TNFα. CD11b^+^ cells were stimulated with **(B)** PMA or **(C)** OVA. Cells were then pulsed with either PD-L1 or CD200 +/- ARL200 with ARL200. Following 48hr incubation, supernatants were analyzed for cytokine production. CD11b cells were pulsed with ARL200, PD-L1, or a combination of ARL200 + PD-L1. **(D)** Cells were immunoaffinity-purified by phospho-peptide enrichment followed by mass-spectroscopy (red, upregulated; green, downregulated). Data were compared for a correlation between the ARL200 vs. PD-L1 and **(E&F)** ARL200 vs. PD-L1 + ARL200 groups.

We next evaluated if ARL200 overpowered the suppressive effects of PD-L1. Splenic CD11b cells were stimulated with PMA or ovalbumin (OVA_257-264_, which contains the antigen SIINFEKL for an immune response). Following exposure to a combination of PD-L1 +/- ARL200 or CD200 +/- ARL200, supernatants were analyzed for cytokine production **(Figure 2b&c)**.

These experiments led us to hypothesize that ARL200 overpowers the inhibitory CD200 and PD-1 ligands. Therefore, we wanted to determine if ARL200 controlled PD-L1-induced signaling pathways. Splenic CD11b cells were isolated from C57BL/6 mice and treated for 5 minutes with ARL200, PD-L1, or a combination of ARL200 + PD-L1. Cells were harvested, and phosphorylated proteins were isolated via pull-down. Treatment groups were normalized to non-treated cells to generate a heat map of 350 results with ≥3-fold change. Significant changes in phosphorylation between ARL200 versus PD-L1 treatment cell groups were noted. Importantly, the phosphorylation profiles of cells pulsed with ARL200+PD-L1 were similar to cells treated with ARL200 alone, indicating the ability of ARL200 to suppress the PD-L1 pathway **(Figure 2d).** Further analyses demonstrated that cells pulsed with ARL200 had a significantly different phosphorylation response compared to those pulsed with PD-L1 (R^2^=0.337). In contrast, the addition of ARL200 with PD-L1, recapitulated the effect of cells pulsed with ARL200 alone (R^2^=0.8412) **(Figure 2e&f)**.

### ARL200 signaling is associated with the downregulation of the inhibitory immune checkpoints through JAK1/3–SHP–STAT–IKKα/β–NFκB

Consistent with the known signaling mechanisms of DAP10&12, we previously reported that ARL200 activates the PI3K–AKT and Ras–MEK–ERK (MAPK) pathways, respectively.^20^ Specifically, we found that ARL200 enhanced DAP10-linked PI3K signaling in splenic CD11b^+^ myeloid cells and that DAP12 signaling pathways involve Janus kinase (JAK)1/3, Src homology 2 domain-containing protein tyrosine phosphatase-2 (SHP-2), and nuclear factor kappa B (NFκB), signaling molecules known to be important for tumor immunity.^37, 20, 38–40^ We further demonstrated that ARL200 signaled through DAP12-associated SYK target. We reported that pSLC76. ERK1/2; and p38MAPK showed an initial increase. In contrast, PI3K and cJUN were significantly increased at later time points, supporting a DAP12-SYK signaling cascade in these myeloid cells.^20^ However, this effect was paralleled by a reduction in key downstream signaling molecules of the PD-1/PD-L1 axis (SHP-1, SHP-2) and a notable decrease in NFκB signaling (NFκB, pIKKα/β) (**Supplemental Figure 1a**). Taken together, ARL200 is signals through specific signaling pathways connected to multiple immune checkpoints.

These observations (**Summarized in Supplemental Figure 1b**) suggest that ARL200 reduces immunosuppressive IC pathways by inhibiting NFκB (or other downstream PI3K/AKT targets) in myeloid cells, supporting that ARL200 elicits multifaceted immune activation. We propose that ARL200 stimulates DAP10-PI3K-AKT and DAP12-SYK-MAPK and other uncharacterized downstream signaling pathways to augment cytokine production, potentially via cJUN. Additionally, crosstalk between the JAK/STAT and NFκB signaling pathways (usually a positive feedback loop) has been reported and is associated with SYK and ZAP70.^41^ This is possibly attributed to the ARL200 inhibition of NFκB signaling, known to upregulate immune checkpoints.^42–48^

Therefore, we evaluated the signaling pathways elicited by ARL200. Interestingly, in addition to SYK and ZAP70, ARL200 downregulates JAK1/3, which is involved in the upregulation of PD-1 and PD-L1 signaling pathways through the upregulation of NFκB.^42–44^ In these studies, CD11b^+^ cells were pulsed with ARL200 and analyzed for the expression levels of various signaling molecules. We discovered that ARL200/CD200AR ligation downregulated JAK1/3–SHP–STAT–IKKα/β–NFκB pathway (**Figure 3a–h**). In addition, we observed that downregulation of CD200R1 and PD1 receptors and their ligands CD200 protein and PD-L1, respectively (**Figure 3i-l**). These studies correlate with our previous inhibition studies, showing that the signaling molecules p38MAPK, JAK1/2, MEK1/2, and JNK1/2 were involved in ARL200-mediated signaling.^20^

**Figure 3.**
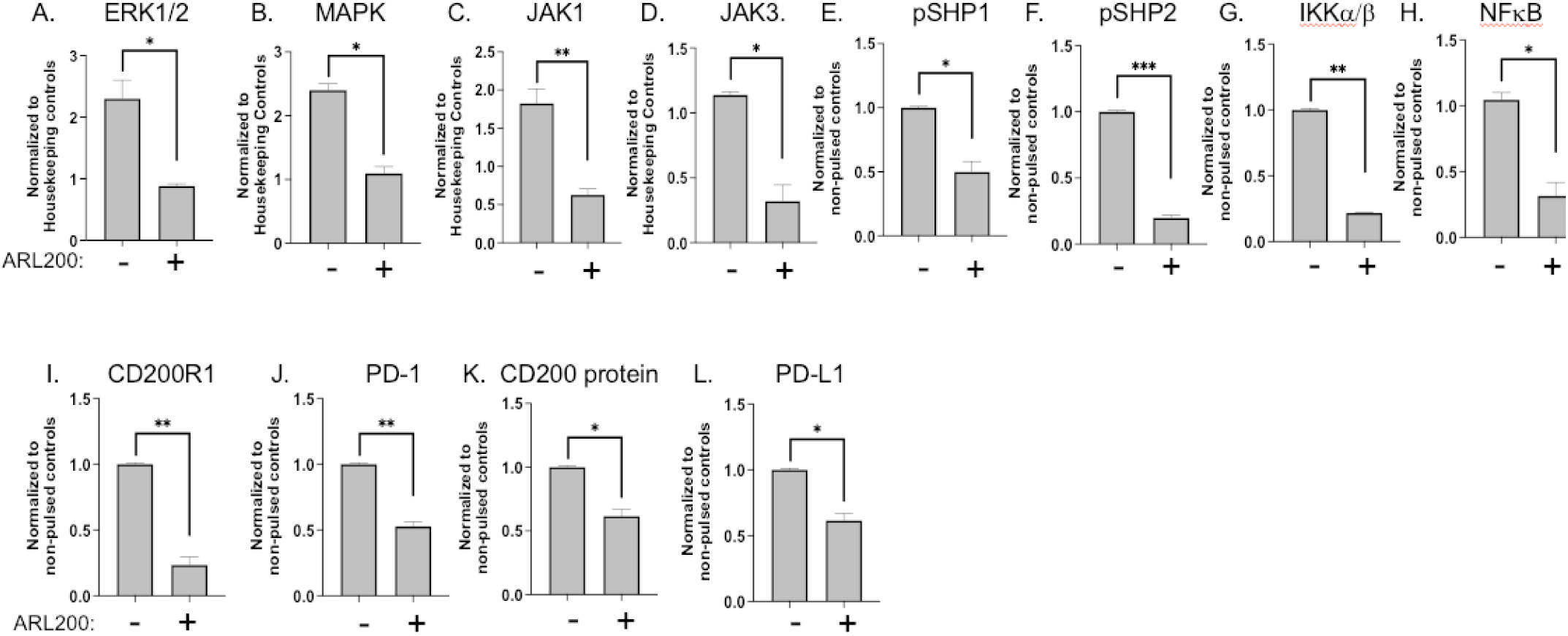
ARL200 regulates PD-1 expression through JAK1/3 and NF-κB expression. CD11b^+^ cells were pulsed with ARL200 and analyzed for (A) ERK1/2, (B) MAPK, (C) JAK1 and (D) JAK3 transcription; (E) SHP1, (F) SHP2 phosphorylation; and (G) IKKα/β, and (H) NF-κB protein expression. JAK1 and JAK3 were normalized to housekeeping genes; all other molecules were normalized to specific non-pulse controls. Error bars, SD; *p < 0.05; **p < 0.01 by multiple comparison test. Representative data from 3 experiments with n=3 /group.

### Downregulation of NCRs by ARL200 translates to a subset of patients with recurrent Glioblastoma

To translate these findings, we initiated a first-in-human trial (NCT04642937) treating adults with recurrent/progressive glioblastoma. Six patients were treated with 3.75 ug/kg of a human sequence of the ARL200 + 1 mg GBM6-AD tumor lysate as a source of tumor antigens (using imiquimod as a topical immune primer as in prior studies of GBM6-AD), both administered subcutaneously **Supplemental Figure 2**).

Three patients (50%) exhibited radiologic responses (termed responders), defined as either >50% response (n=1) or stable disease (n=2) determined by MRI. The remaining three patients lacked any evidence of radiologic response (termed non-responders). We observed that responders presented with decreased levels of ICs and reduction in the numbers of suppressive regulatory cells post-treatment, in contrast to the non-responders (**Figure 4a-d**). In addition, we observed enhanced immune activation in responders as determined by flow cytometry and bulkRNA (**Figure 4e-i)**. To determine if we elicited an antigen-specific memory response, purified patient PBMCs were pulsed with GBM6-AD tumor lysates, and supernatants were analyzed for TNFα and IL-17 (**Figure 4j&k)**. These studies suggest that the mechanism of the ARL200 translated to humans and is potentially responsible for the extended survival observed in the responder population (**Figure 4l)**. Although these experiments only involved a small patient population and only suggested potential efficacy, we have initiated a study treating children and young adults with recurrent high-grade glioma and primary diffuse midline glioma. In both trials, we have not observed any drug-related serious adverse effects.

**Figure 4.**
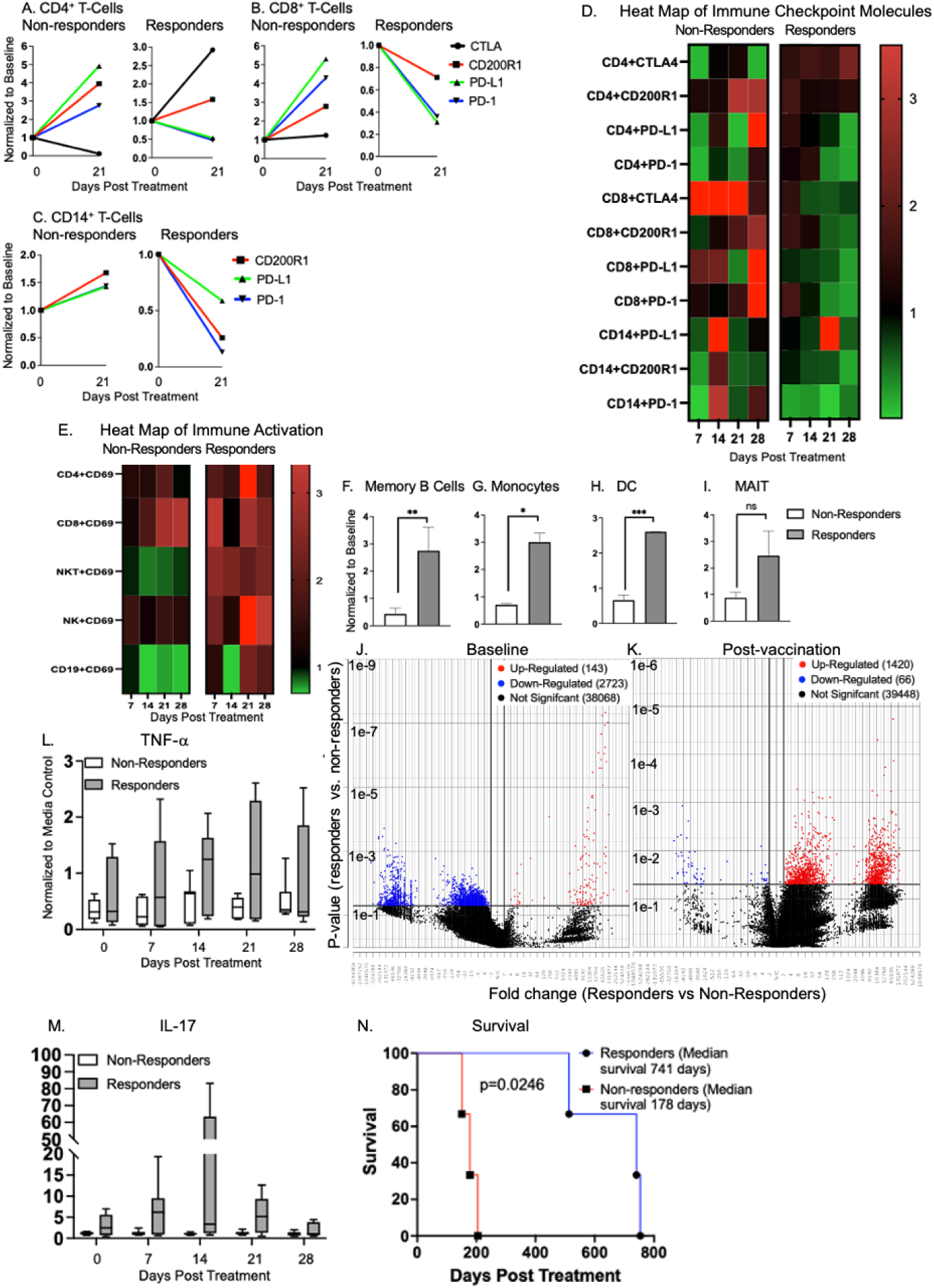
ARL200 activates the immune system and downregulates regulatory and IC molecules. In a Phase I dose-escalation trial, 6 patients received 3.25 µg/kg ARL200 + TL for 4 consecutive weeks (induction period, Supplemental Figure 2). Whole blood was analyzed pre-and post-treatment from clinically responding and non-clinically responding patients. Absolute numbers of immune checkpoint receptors and ligands were calculated on **(A)** CD4 T-cells, **(B)** CD8 T-cells and **(C)** CD14 cells. Heat maps of absolute numbers normalized to baseline were derived for **(D)** immune checkpoint molecules and **(E)** immune activation. Purified lymphocytes were analyzed by BulkRNA for **(F)** MALT, **(G)** Monocytes, **(H)** dendritic cells (DC), and **(I)** MAIT. **(J&K)** volcano plots from baseline and one week post vaccination. Freshly purified lymphocytes were pulsed with GBM6-AD tumor lysates, incubated for 48hrs and analyzed for **(L)** TNF-a and **(M)** IL-17. Cells were normalized to non-pulsed control. **(N)** Patients were followed for overall survival.

## Discussion

Our novel peptide ligand, ARL200 represents a pivotal advancement through unique and potent mechanisms that stimulate an antitumor response through simultaneously downregulating multiple immune checkpoints CD200R1, PD-1, and CTLA-4 and their suppressive ligands CD200 protein and PD-L1.^16–19, 49^ The ability to modulate multiple immune checkpoints with this single peptide is a groundbreaking discovery;^15^ it is well accepted that targeting multiple immune checkpoints results in enhanced survival.^15^ This is not limited to current immune checkpoint antibodies; others have reported that the use of signaling inhibitors, such as mitogen-activated protein kinase (MAPK) signaling, was associated with an enhanced clinical response, specifically in glioblastoma, where even combination immune checkpoint blockade fails.^4, 50, 51^

To this end, it is critically important to understand the complex immune regulatory mechanisms of immune checkpoint receptors as well as the disease-specific immune milieu. In addition to the complex pathways controlling NCRs, we posit that signaling pathways elicited by different CD200AR complexes drive alternative immune responses. The CD200 activation receptor is a complex of multiple CD200ARs.^15^ It is known that there are two activation receptors (CD200ARLa&b) in humans and four in mice (CD200AR2-5), however, CD200AR5 is a pseudogene and not expressed.^25, 27, 52^ These activation receptors are derived from the CD200R1, likely due to evolutionary gene duplication events,^52^ and share 83%, 39%, 84%, and 91% homology between the inhibitory CD200R1 and CD200AR2, CD200AR3, CD200AR4, and CD200AR5 respectively.^52, 53^

Our murine CD200AR knock-out studies determined that single CD200ARs do not bind the ARL200. Rather, a complex of two CD200ARs is required,^20^ and the combination of the different CD200AR2, CD200AR3, & CD200AR4 complexes elicit alternative immune resposnes.^19, 20^ These complexes are important for the targeting of different tumor locations. We previously reported that the ARL200 sequence described in this manuscript binds to a specific CD200AR complex, eliciting a precise immune response.^19^ In contrast to the ARL200 (termed ARL200a for the remainder of this manuscript), we observed cells pulsed with a second ARL200 sequence (ARL200b) had a different immune response (**Supplemental Figure 3a).** Similarly, human CD14^+^ cells pulsed with humanized peptides exhibited distinct signaling and chemokine profiles **(Supplemental Figure 3b).**^16, 19^ We posit that these different responses result in alternative anti-tumor responses. We previously reported that ARL200a (the ARL200 used in all the data reported above) demonstrated efficacy in glioma, but low efficacy in breast carcinoma models.^16^ In contrast, the ARL200b sequence demonstrated high efficacy in breast carcinoma models but failed in glioma models.^16^ We repeated the breast cancer studies using breast organoids implanted in the murine fat pad, confirming consistent peptide activity for ARL200b but losing durability with ARL200a.^54^ Remarkably, when mice were rechallenged by injection of organoids into the contralateral fat pad following regression, both peptides demonstrated a robust memory response compared to control (ARL200b**; Supplemental Figure 3c&d).** To determine if this was tumor specific or the difference was due to CNS or peripheral locations, breast cancer organoids were introduced to the cerebellum and mice were then treated with ARL200a or ARL200b. In contrast to non-CNS breast tumors, ARL200a was more effective against the breast cancer organoids in the cerebellum than ARL200b **(Supplemental Figure 3e-g)**.

We know that the ARL200a primarily binds to a combination of CD200AR2 & CD200AR3 receptors,^19^ we believe that the different immune responses are due to the ARL200b binding to the CD200AR2 & CD200AR4 or CD200AR3 & CD200AR4 complexes, activating different signaling pathways. This supports our working hypothesis that ARL200a and ARL200b signal through alternative signaling pathways. Our studies demonstrate that the ARL200a signals through the JAK1/3–SHP–STAT–IKKα/β–NFκB as demonstrated in **Figure 3**; as recent preliminary studies showed that ARL200b does not signal through pSHP1/2, IKKα/β, or NFκB, but both ARL200a and ARL200b peptides continue to downregulate multiple NCRs **(Supplemental Figure 4a-g)**.

In conclusion, we have developed a next-generation drug that targets multiple NCR through common signaling molecules like protein tyrosine phosphatase and non-receptor type 6 (SHP-1/2), both known targets for cancer immunotherapy.^5^ This mechanism allows clinicians to eliminate the inhibitory effects of multiple immune checkpoints with a single drug, eliminating potential serious adverse effects. In addition to the elimination of NCRs, we are potentially capable of controlling the immune response required for different tumors, depending on the type and location of the tumor in each patient.

## Data Availability

All data produced in are in the manuscript

## Acknowledgements

We would like to thank Itay Raphael and Franscisco Puerta for their assistance on the manuscript. We would like to thank Randy Shaver Cancer Research and Community Fund, Children Cancer Research Fund and Humor to Fight the Tumor and OX2 Therapeutics for their support. In addition, this work was supported by the Department of Defense Congressionally Directed Medical Research Programs under award number HT9425-25-1-0244 The views expressed are those of the authors and do not necessarily reflect the official policy or position of the U.S. Government.

## Authors contribution

Maghna Saxena contributed to experiments shown in figures 1, 3, and supplemental figures 1&4. She also contributed to writing and review of final manuscript. Elisabeth Ampudia-Mesias contributed to experiments 1, 2 and 3. Sumant Dhawan contributed to figure 4, writing and review of the manuscript. Gary Kohanbash contributed to bulkRNA analysis shown in figure 4, design of Supplemental Figure 1, writing and review of the manuscript. Stephen C. Frederico contributed to the writing, format and submission of the manuscript. Xiaoqing Cheng and Ron Bose contributed to supplementary figure 3, writing and review of the manuscript. Elisabeth Neil was the PI of the clinical trial reported in figure 4 and Supplemental Figure 2 and reviewed the final manuscript. Christopher L Moertel contributed to design of all figures, writing and review of manuscript. Michael Olin contributed to all aspects of the experimental designs, analysis and manuscript writing and review.

Data will be made available upon request.

---

**Supplemental Figure 1.**
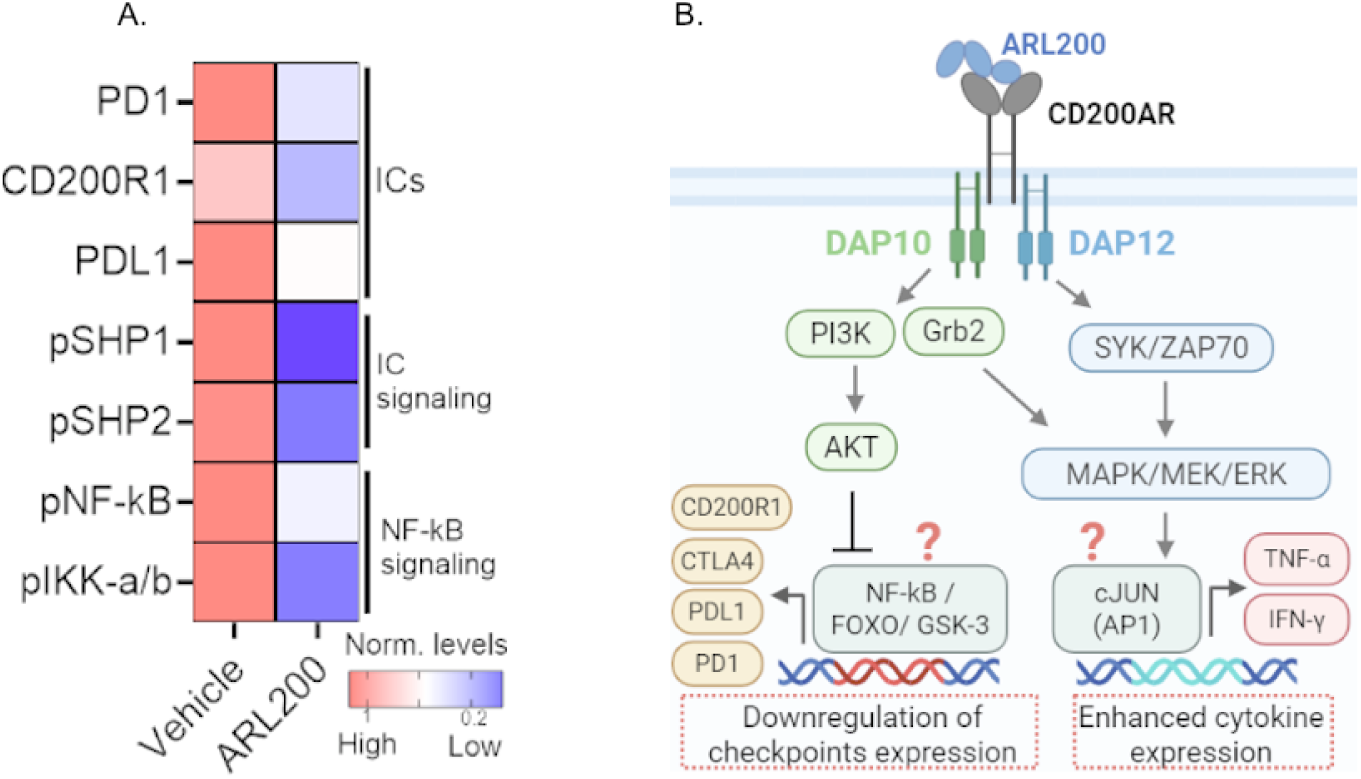
ARL200 inhibits immune checkpoints molecules and NF-kB signaling. CD11b^+^ myeloid cells isolated from WT mice were pulsed with 10 µM (0.2 µg) ARL200. Cells were isolated and analyzed by western blot. (**A**) Heatmap showing quantified protein levels of immune checkpoints (PD-1, PD-L1, and CD200R1; top), their signaling molecules SHP-1 and SHP2 (middle), and inhibition of the NF-κB pathway (bottom) in CD11b^+^ cells stimulated with ARL200 relative to vehicle. (**B**) Hypothesizes of ARL200 intracellular signaling. DAP10 vs DAP12 expression levels between cell types impacting specific responses. Putatively, DAP10 and DAP12 are relatively higher in myeloid cells and T-cells, respectively. Additional signaling downstream of the cytokines and immune checkpoints are not shown but elaborated on in the text and in Figure 3. Error bars, SD; *p < 0.05; **p < 0.01 by multiple comparison test. Representative data from 3 experiments with n=3 /group.

**Supplementary figure 2.**
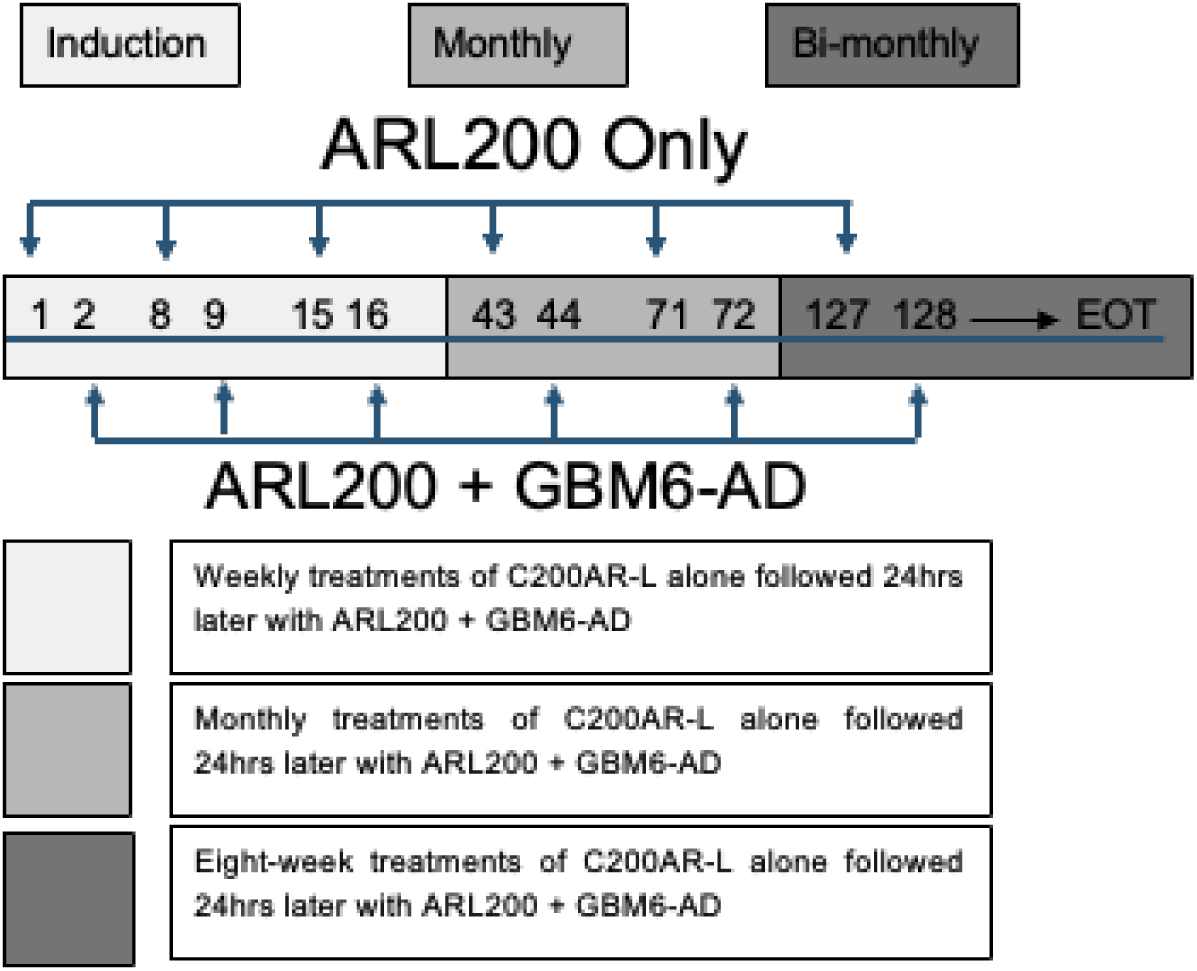
ALR200 and GBM6-AD vaccine administered as follows: On day 1 and 2 of each treatment series, hP1A8 at assigned dose by intradermal injection, divided between two supraclavicular sites, after application of topical imiquimod. On day 2 only, GBM6-AD is administered at a dose of 1 mg lysate b intradermal injection, divided between the same sites, within 30 minutes following ALR200 injections until end of treatment (EOT)

**Supplemental Figure 3.**
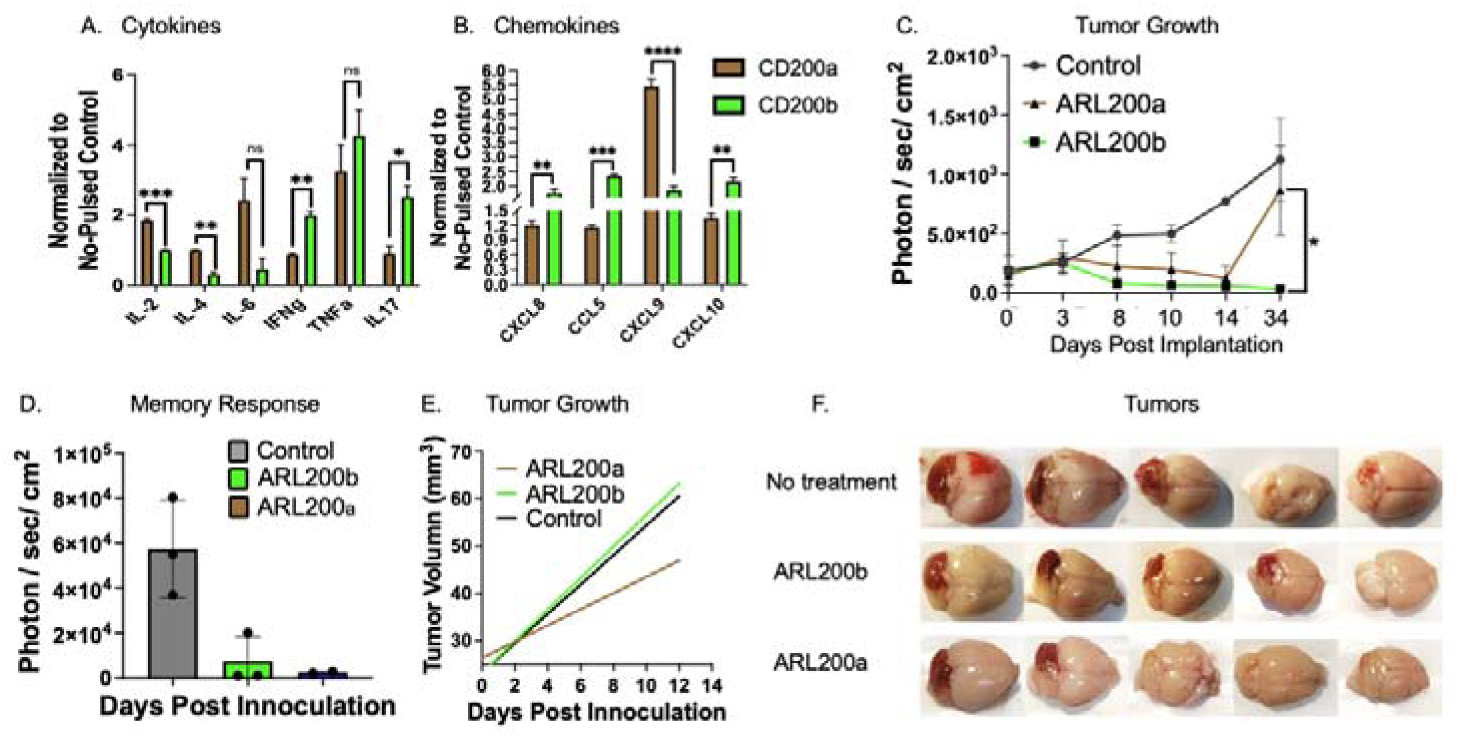
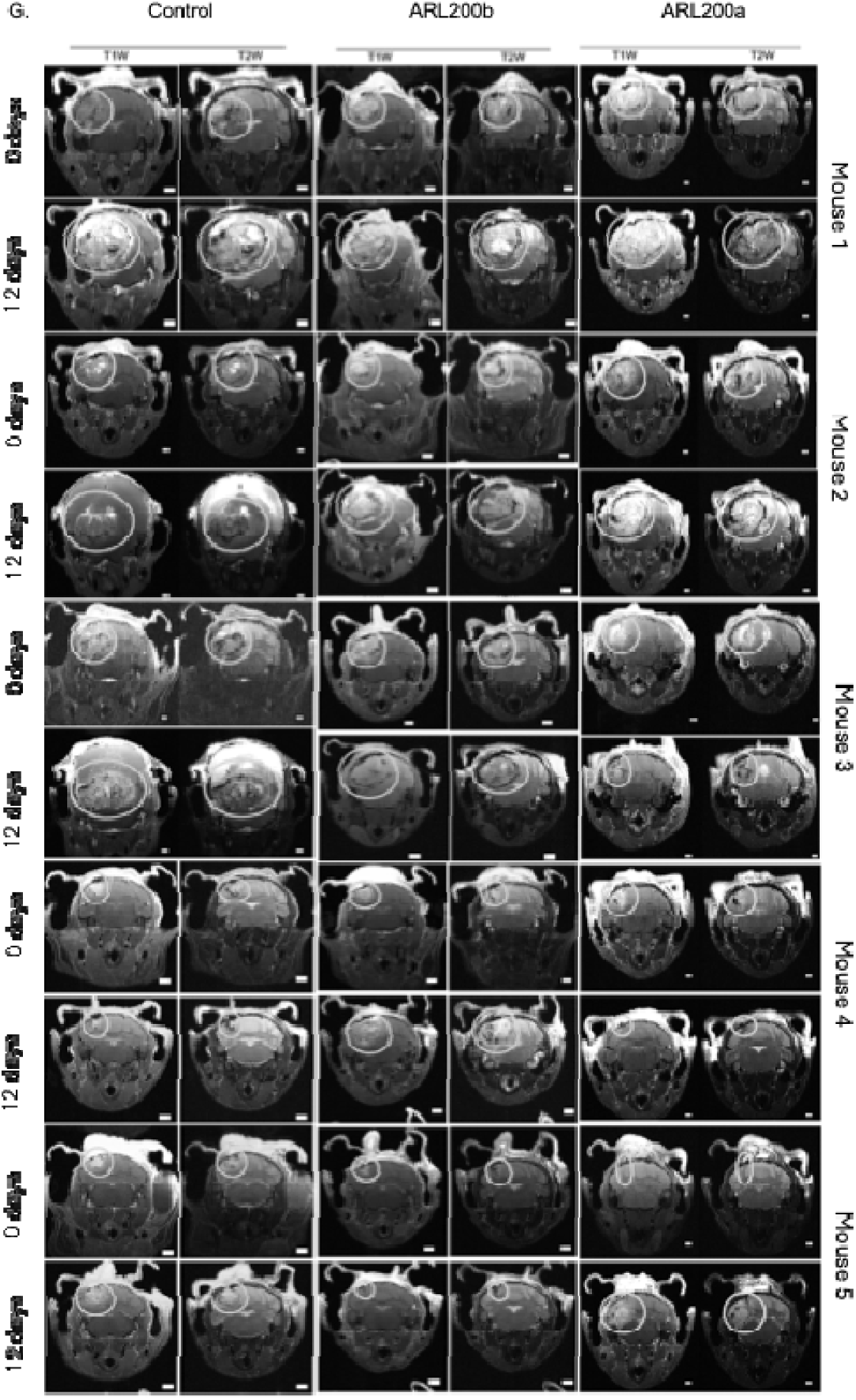
ARL200 activity differs in primary breast cancer and CNS metastases models. **(A)** Cytokines profiling in CD11b+ cells pulsed with ARL200 treatment after normalization. **(B)** Chemokines profiling in human CD14+ cells pulsed with ARL200 treatment after normalization. **(C)** BLI intensity measurements of HP organoid transplanted tumor growth on luciferase signal on indicated days post-drug treatment. N>=3 per arm. **(D)** BLI intensity measurements of HP organoid transplanted tumor growth on luciferase signal on indicated days post-inoculation for memory response testing. N>=3 per arm. **(E)** Tumor volume measurements of HP organoid transplanted tumor growth on indicated days post-inoculation for memory response testing. N>=3 per arm. **(F)** Brightfield images of dissected cerebellum tumor from control and anti-ARL200 groups. **(G)** The representative T1W and T2W images of MRI showed the cerebellar tumor volume on days and 12 days post-treatment from control and anti-ARL200 groups. 5000 cells of *HER2^V777L^*; *PIK3CA*^H1047R^ (HP) tumor organoid were injected into the mouse cerebellum for modeling as described in the methods.

**Supplemental Figure 4.**
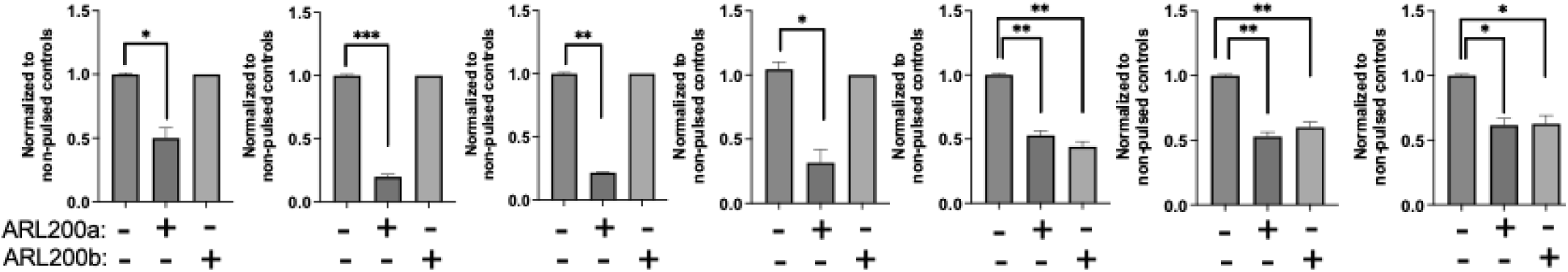
ALR200a and ARL200b signal through an alternat signaling pathway. CD11b^+^ cells were pulsed with ARL200 and analyzed for (A) SHP1, (B) SHP2, (C) IKKα/β, (D) NFκB and (E) CD200R1, (F) PD-1 and (G) PD-L1 protein expression by western analysis. All molecules were normalized to specific non-pulse controls. Error bars, SD; *p < 0.05; **p < 0.01 by multiple comparison test. Representative data from 3 experiments with n=3/group.

## References

1. Abbott M, Ustoyev Y. Cancer and the Immune System: The History and Background of Immunotherapy. Semin Oncol Nurs. 2019;35: 150923.

2. Madden K, Kasler MK. Immune Checkpoint Inhibitors in Lung Cancer and Melanoma. Semin Oncol Nurs. 2019;35: 150932.

3. Borggrewe M, Kooistra SM, Noelle RJ, Eggen BJL, Laman JD. Exploring the VISTA of microglia: immune checkpoints in CNS inflammation. J Mol Med (Berl). 2020;98: 1415–1430.

4. Arafat Hossain M. A comprehensive review of immune checkpoint inhibitors for cancer treatment. Int Immunopharmacol. 2024;143: 113365.

5. Watson HA, Wehenkel S, Matthews J, Ager A. SHP-1: the next checkpoint target for cancer immunotherapy? Biochem Soc Trans. 2016;44: 356–362.

6. Curdy N, Lanvin O, Laurent C, Fournié JJ, Franchini DM. Regulatory Mechanisms of Inhibitory Immune Checkpoint Receptors Expression. Trends Cell Biol. 2019;29: 777–790.

7. Salik B, Smyth MJ, Nakamura K. Targeting immune checkpoints in hematological malignancies. J Hematol Oncol. 2020;13: 111.

8. Darvin P, Toor SM, Sasidharan Nair V, Elkord E. Immune checkpoint inhibitors: recent progress and potential biomarkers. Exp Mol Med. 2018;50: 1–11.

9. Tivol EA, Borriello F, Schweitzer AN, Lynch WP, Bluestone JA, Sharpe AH. Loss of CTLA-4 leads to massive lymphoproliferation and fatal multiorgan tissue destruction, revealing a critical negative regulatory role of CTLA-4. Immunity. 1995;3: 541–547.

10. Francisco LM, Sage PT, Sharpe AH. The PD-1 pathway in tolerance and autoimmunity. Immunol Rev. 2010;236: 219–242.

11. Feng Y, Roy A, Masson E, Chen TT, Humphrey R, Weber JS. Exposure-response relationships of the efficacy and safety of ipilimumab in patients with advanced melanoma. Clin Cancer Res. 2013;19: 3977–3986.

12. Ho AK, Ho AM, Cooksley T, Nguyen G, Erb J, Mizubuti GB. Immune-Related Adverse Events Associated With Immune Checkpoint Inhibitor Therapy. Anesth Analg. 2021;132: 374–383.

13. Dougan M, Luoma AM, Dougan SK, Wucherpfennig KW. Understanding and treating the inflammatory adverse events of cancer immunotherapy. Cell. 2021;184: 1575–1588.

14. Pauken KE, Dougan M, Rose NR, Lichtman AH, Sharpe AH. Adverse Events Following Cancer Immunotherapy: Obstacles and Opportunities. Trends Immunol. 2019;40: 511–523.

15. Kottschade LA. The Future of Immunotherapy in the Treatment of Cancer. Semin Oncol Nurs. 2019;35: 150934.

16. Moertel CL, Xia J, LaRue R, et al. CD200 in CNS tumor-induced immunosuppression: the role for CD200 pathway blockade in targeted immunotherapy. J Immunother Cancer. 2014;2: 46.

17. Moertel C, Martinez-Puerta F, Pluhar GGE, Castro MG, Olin M. CD200AR-L: mechanism of action and preclinical and clinical insights for treating high-grade brain tumors. Expert Opin Investig Drugs. 2022;31: 875–879.

18. Xiong Z, Ampudia-Mesias E, Shaver R, Horbinski CM, Moertel CL, Olin MR. Tumor-derived vaccines containing CD200 inhibit immune activation: implications for immunotherapy. Immunotherapy. 2016;8: 1059–1071.

19. Xiong Z, Ampudia Mesias E, Pluhar GE, et al. CD200 Checkpoint Reversal: A Novel Approach to Immunotherapy. Clin Cancer Res. 2019;26: 232–241.

20. Ampudia-Mesias E, Puerta-Marttinez F, Bridges M, et al. CD200 Immune-Checkpoint Peptide Elicits an Anti-glioma Response Through the DAP10 Signaling Pathway. Neurotherapeutics. 2021;18: 1980–1994.

21. Kretz-Rommel A, Qin F, Dakappagari N, et al. CD200 expression on tumor cells suppresses antitumor immunity: new approaches to cancer immunotherapy. J Immunol. 2007;178: 5595–5605.

22. Coles SJ, Wang EC, Man S, et al. CD200 expression suppresses natural killer cell function and directly inhibits patient anti-tumor response in acute myeloid leukemia. Leukemia. 2011;25: 792–799.

23. Raphael I, Mujeeb AA, Ampudia-Mesias E, et al. CD200 depletion in glioma enhances antitumor immunity and induces tumor rejection. bioRxiv. 2024: 2024.2009.2008.611922.

24. Kretz-Rommel A, Qin F, Dakappagari N, et al. CD200 expression on tumor cells suppresses antitumor immunity: new approaches to cancer immunotherapy. J Immunol. 2007;178: 5595–5605.

25. Gorczynski R, Chen Z, Kai Y, Lee L, Wong S, Marsden PA. CD200 is a ligand for all members of the CD200R family of immunoregulatory molecules. J Immunol. 2004;172: 7744–7749.

26. Gorczynski RM, Chen Z, Diao J, et al. Breast cancer cell CD200 expression regulates immune response to EMT6 tumor cells in mice. Breast Cancer Res Treat. 2010;123: 405–415.

27. Voehringer D, Rosen DB, Lanier LL, Locksley RM. CD200 receptor family members represent novel DAP12-associated activating receptors on basophils and mast cells. J Biol Chem. 2004;279: 54117–54123.

28. Pallasch CP, Ulbrich S, Brinker R, Hallek M, Uger RA, CM W. Disruption of T cell suppression in chronic lymphocytic leukemia by CD200 blockade. Leuk Res. 2009;33: 460–464.

29. Stokes MP, Farnsworth CL, Gu H, et al. Complementary PTM Profiling of Drug Response in Human Gastric Carcinoma by Immunoaffinity and IMAC Methods with Total Proteome Analysis. Proteomes. 2015;3: 160–183.

30. Possemato A, Paulo J, Mulhern D, Guo A, Gygi S, Beausoleil S. Multiplexed Phosphoproteomic Profiling Using Titanium Dioxide and Immunoaffinity Enrichments Reveals Complementary Phosphorylation Events. J Proteome Res. 2017;16: 1506–1514.

31. MacLean B, Tomazela DM, Shulman N, et al. Skyline: an open source document editor for creating and analyzing targeted proteomics experiments. Bioinformatics. 2010;26: 966–968.

32. Kretz-Rommel A, Qin F, Dakappagari N, Cofiell R, Faas SJ, Bowdish KS. Blockade of CD200 in the presence or absence of antibody effector function: implications for anti-CD200 therapy. J Immunol. 2008;180: 699–705.

33. Mahadevan D, Lanasa MC, Farber C, et al. Phase I study of samalizumab in chronic lymphocytic leukemia and multiple myeloma: blockade of the immune checkpoint CD200. J Immunother Cancer. 2019;7: 227.

34. Jens M, Chemnitz R, Parry K, Nichols, Jube C, Riley J. SHP-1 and SHP-2 Associate with Immunoreceptor Tyrosine-Based Switch Motif of Programmed Death 1 upon Primary Human T Cell Stimulation, but Only Receptor Ligation Prevents T Cell Activation. J Immunol. 2014;173: 945–954.

35. Zhang S, Cherwinski H, Sedgwick JD, Phillips JH. Molecular mechanisms of CD200 inhibition of mast cell activation. J Immunol. 2004;173: 6786–6793.

36. Coles SJ, Gilmour MN, Reid R, et al. The immunosuppressive ligands PD-L1 and CD200 are linked in AML T-cell immunosuppression: identification of a new immunotherapeutic synapse. Leukemia. 2015.

37. Lorenz U. SHP-1 and SHP-2 in T cells: two phosphatases functioning at many levels. Immunol Rev. 2009;228: 342–359.

38. Brooks AJ, Putoczki T. JAK-STAT Signalling Pathway in Cancer. Cancers (Basel). 2020;12.

39. Chan L, Wang S, Hung M. IL-6/JAK1 pathway drives PD-L1 Y112 phosphorylation to promote cancer immune evasion. The Journal of Clinical Investigation. 2019;129: 3324–3338.

40. Zhou S, Wang B, Wei Y, et al. PD-1 inhibitor combined with Docetaxel exerts synergistic anti-prostate cancer effect in mice by down-regulating the expression of PI3K/AKT/NFKB-P65/PD-L1 signaling pathway. Cancer Biomark. 2024;40: 47–59.

41. Hu X, li J, Fu M, Zhao X, Wang W. The JAK/STAT signaling pathway: from bench to clinic. Signal Transduction and Targeted Therapy. 2021;6: 402.

42. Mirzaei R, Gordon A, Zemp FJ, et al. PD-1 independent of PD-L1 ligation promotes glioblastoma growth through the NFkappaB pathway. Sci Adv. 2021;7: eabh2148.

43. Sharpe AH, Pauken KE. The diverse functions of the PD1 inhibitory pathway. Nat Rev Immunol. 2018;18: 153–167.

44. Rolvering C, Zimmer AD, Ginolhac A, et al. The PD-L1- and IL6-mediated dampening of the IL27/STAT1 anticancer responses are prevented by alpha-PD-L1 or alpha-IL6 antibodies. J Leukoc Biol. 2018;104: 969–985.

45. Yi YS, Son YJ, Ryou C, Sung GH, Kim JH, Cho JY. Functional roles of Syk in macrophage-mediated inflammatory responses. Mediators Inflamm. 2014;2014: 270302.

46. Beard A, Trotter SC. JAK 1-3 inhibitors and TYK-2 inhibitors in dermatology: Practical pearls for the primary care physician. J Family Med Prim Care. 2024;13: 4128–4134.

47. Demir M, Cizmecioglu O. ZAP70 Activation Compensates for Loss of Class IA PI3K Isoforms Through Activation of the JAK-STAT3 Pathway. Cancer Diagn Progn. 2022;2: 391–404.

48. Wang H, Kadlecek TA, Au-Yeung BB, et al. ZAP-70: an essential kinase in T-cell signaling. Cold Spring Harb Perspect Biol. 2010;2: a002279.

49. Choe D, Choi D. Cancel cancer: The immunotherapeutic potential of CD200/CD200R blockade. Frontiers. 2023;13.

50. Zhao J, Chen AX, Gartrell RD, et al. Immune and genomic correlates of response to anti-PD-1 immunotherapy in glioblastoma. Nat Med. 2019;25: 462–469.

51. Arrieta VA, Chen AX, Kane JR, et al. ERK1/2 phosphorylation predicts survival following anti-PD-1 immunotherapy in recurrent glioblastoma. Nat Cancer. 2021;2: 1372–1386.

52. Wright GJ CH, Foster-Cuevas M, Brooke G, Puklavec MJ, Bigler M, Song Y, Jenmalm M, Gorman D, McClanahan T, Liu MR, Brown MH, Sedgwick JD, Phillips JH, Barclay AN. Characterization of the CD200 receptor family in mice and humans and their interactions with CD200. J Immunol. 2003;171.

53. Hatherley D, Barclay A. The CD200 and CD200 receptor cell surface proteins interact through their N-terminal immunoglobulin-like domains. Eur J Immunol. 2004;34: 1688–1694.

54. Cheng X, Sun Y, Highkin M, et al. Breast Cancer Mutations HER2V777L and PIK3CAH1047R Activate the p21-CDK4/6-Cyclin D1 Axis to Drive Tumorigenesis and Drug Resistance. Cancer Res. 2023;83: 2839–2857.

